# Association between visit-to-visit lipid variability and incident cancer: a population-based cohort study

**DOI:** 10.1101/2022.09.04.22279557

**Authors:** Jeffrey Shi Kai Chan, Danish Iltaf Satti, Yan Hiu Athena Lee, Khalid Bin Waleed, Pias Tang, Gauranga Mahalwar, Abdul Mannan Khan Minhas, Leonardo Roever, Giuseppe Biondi-Zoccai, Fung Ping Leung, Wing Tak Wong, Tong Liu, Jiandong Zhou, Gary Tse

**Affiliations:** Family Medicine Research Unit, Cardiovascular Analytics Group, United Kingdom – Hong Kong – China collaboration; Department of Cardiology, St George’s University Hospital NHS Foundation Trust, London, United Kingdom; Department of Internal Medicine, Cleveland Clinic Akron General, Akron, Ohio, United States of America; Department of Medicine, Forrest General Hospital, Hattiesburg, Mississippi, United States of America; Departamento de Pesquisa Clinica, Universidade Federal de Uberlandia, Uberlandia, MG, Brazil; Department of Medical-Surgical Sciences and Biotechnologies, Sapienza University of Rome, Latina, Italy; Mediterranea Cardiocentro, Napoli, Italy; School of Life Sciences, The Chinese University of Hong Kong, Hong Kong, China; Tianjin Key Laboratory of Ionic-Molecular Function of Cardiovascular Disease, Department of Cardiology, Tianjin Institute of Cardiology, Second Hospital of Tianjin Medical University, Tianjin 300211, China; Nuffield Department of Medicine, University of Oxford, Oxford, United Kingdom; Kent and Medway Medical School, Canterbury, Kent, CT2 7NT, United Kingdom; Epidemiology Research Unit, Cardiovascular Analytics Group, United Kingdom – Hong Kong – China collaboration

**Author notes:** Corresponding authors: *Dr Jiandong Zhou, PhD*, Nuffield Department of Medicine, University of Oxford, Oxford, United Kingdom, Family Medicine Research Unit, Cardiovascular Analytics Group, United Kingdom – Hong Kong – China collaboration, *Dr Gary Tse, MD PhD FRCP FFPH*, Kent and Medway Medical School, Canterbury, Kent, CT2 7NT, United Kingdom, Epidemiology Research Unit, Cardiovascular Analytics Group, Hong Kong – United Kingdom – China collaboration, Tianjin Key Laboratory of Ionic-Molecular Function of Cardiovascular Disease, Department of Cardiology, Tianjin Institute of Cardiology, Second Hospital of Tianjin Medical University, Tianjin 300211, China. Co-first authors (Jeffrey Shi Kai Chan, Danish Iltaf Satti). **Funding:** This work was funded by the Tianjin Key Medical Discipline (Specialty) Construction Project (Project number: TJYXZDXK-029A).

**Keywords:** cholesterol, variability, family medicine, preventive cardiology, cancer

## Abstract

**Background:** Dyslipidaemia is associated with increased cancer risk. However, the prognostic value of visit-to-visit lipid variability (VVLV) is unexplored in this regard.

**Objective:** To investigate the associations between VVLV and the risk of incident cancer.

**Design:** Retrospective cohort study.

**Setting:** Family medicine clinics.

**Patients:** Adults attending a family medicine clinic in Hong Kong during 2000-2003, excluding those with <3 tests for low-density lipoprotein cholesterol (LDL-C), high-density lipoprotein cholesterol (HDL-C), triglycerides, and total cholesterol (TC) each, those with prior cancer diagnosis, and those with <1 year of follow-up.

**Measurements:** Visit-to-visit LDL-C, HDL-C, TC, and triglycerides variabilities were measured by the coefficient of variation (CV). Patients were followed up until 31^st^ December 2019 for the primary outcome of incident cancer.

**Results:** Altogether, 69,186 patients were included (26,679 males (38.6%); mean age 60±13 years; mean follow-up 16±3 years); 7958 patients (11.5%) had incident cancer. Higher variability of LDL-C, HDL-C, TC, and TG was associated with higher risk of incident cancer. Patients in the third tercile of the CV of LDL-C (adjusted hazard ratio (aHR) against first tercile 1.06 [1.00, 1.12], p=0.049), HDL-C (aHR 1.37 [1.29, 1.44], p<0.001), TC (aHR 1.10 [1.04, 1.17], p=0.001), and TG (aHR 1.11 [1.06, 1.18], p<0.001) had the highest risks of incident cancer. Among these, only HDL-C variability remained associated with the risk of incident cancer in users of statins/fibrates.

**Limitations:** Due to the observational nature of this study, there may be residual and unmeasured confounders. Patient data could not be individually adjudicated, implying that coding errors may be possible.

**Conclusion:** Higher VVLV was associated with significantly higher long-term risks of incident cancer. VVLV may be a clinically useful tool for cancer risk stratification.

## Introduction

Cancer is one of the leading causes of mortality worldwide, with an estimated 19.3 million new cases and nearly 10 million cancer deaths globally in 2020 (1). Being a major global health burden, it is associated with significant mortality, morbidity, and healthcare expenditures (1,2). Therefore, there is a need to identify new biomarkers to identify patients at heightened risks of cancer, in order to facilitate its prevention and treatment.

Many studies have explored risk factors of incident cancer. Of these, dyslipidaemia is considered to be an important modifiable risk factor for cancer. Preclinical evidence suggested that altered lipid metabolism may lead to carcinogenesis, invasion, and metastasis via multiple signalling pathways (3). Additionally, altered levels of serum lipids have been observed in several malignancies (4,5). A prospective analysis of middle-aged and older women demonstrated that abnormal lipid levels were associated with various types of cancers, with lung and colorectal cancer showing particularly strong associations (6), while others have demonstrated a potential link between altered high-density lipoprotein cholesterol (HDL-C) levels and non-Hodgkin lymphoma (7). Nonetheless, the vast majority of studies have focused on lipid levels as mean levels or point estimates, despite lipid levels being known to display significant intra-personal variations on repeated testing (8,9). This variation on repeated testing, most commonly observed when a patient is tested for lipid levels at different clinic visits, has been coined *visit-to-visit lipid variability* (VVLV).

There has been increased interest in VVLV over recent years, particularly its role as a surrogate marker for vascular disease (10). Recent epidemiological research has demonstrated that such variability is associated with numerous adverse clinical events, such as heart failure (11), myocardial infarction (12,13), stroke (13), sudden cardiac death (14), and mortality (11,15). However, its association with incident cancer has been relatively unexplored, despite the aforementioned links between lipid metabolism and incident cancer. As such, this study aimed to investigate the associations between VVLV and incident cancer in the general population.

## Methods

This study was conducted in accordance with the Declaration of Helsinki and the Strengthening the Reporting of Observational Studies in Epidemiology guideline (16), and approved by the Joint Chinese University of Hong Kong– New Territories East Cluster Clinical Research Ethics Committee. The need for individual consent was waived as only retrospective, deidentified data were used. All data were available on reasonable request to the corresponding author.

### Data source

Data were retrieved from the Clinical Data Analysis and Reporting System (CDARS), a deidentified administrative electronic medical database that automatically records basic demographics, diagnoses, selected laboratory tests, and selected medical procedures of all patients attending public healthcare institutions in Hong Kong. Given that these institutions cover the entire territory of Hong Kong and have been estimated to serve 90% of the Hong Kong population (17), CDARS is thus a population-based database and is the most representative electronic medical database available in Hong Kong. Diagnoses in CDARS were coded using the International Classification of Diseases, Ninth revision (ICD-9) codes regardless of the time of entry, as ICD-10 codes have not been implemented in CDARS to date. Mortality data were retrieved from the linked Hong Kong Death Registry, a governmental registry which records mortality data of all Hong Kong citizens. CDARS and the linked Hong Kong Death Registry have been used extensively in prior studies, and have been shown to have good coding accuracy (18–22).

### Patient population

Patients aged 18 years old or above who attended a family medicine clinic in Hong Kong during 1^st^ Jan 2000 to 31^st^ December 2003 were included. Those who had less than three test results for low-density lipoprotein cholesterol (LDL-C), high-density lipoprotein cholesterol (HDL-C), triglycerides, and total cholesterol each, those who had any prior diagnosis of cancer, and those with less than one year of follow-up were excluded.

### Follow-up and outcomes

All patients were followed up until 31^st^ December 2019. The outcome of interest was incident cancer.

### Data collection and definitions

Age, sex, comorbid conditions (hypertension, heart failure, atrial fibrillation, chronic kidney disease, myocardial infarction, ischaemic heart disease, peripheral vascular disease, stroke or transient ischaemic attack, diabetes mellitus, and dyslipidaemia; the corresponding ICD-9 codes are summarized in **Supplementary Table 1**), medication prescriptions (angiotensin-converting enzyme inhibitors or angiotensin receptor blockers, beta-blockers, calcium channel blockers, statins or fibrates, antiplatelets, metformin, sulphonylureas, and insulins), and total cholesterol, LDL-C, HDL-C, and total cholesterol levels at baseline and during follow-up were recorded. Records of dyslipidaemia diagnoses were supplemented by the use of any lipid-lowering medications, a minimum total cholesterol level ≥5.2 mmol/L, a minimum LDL-C level ≥3.4 mmol/L, a maximum HDL-C level <1.0 mmol/L, or a minimum triglycerides level ≥1.7 mmol/L (23). Selected diagnoses were supplemented by medication prescriptions and/or laboratory tests: the diagnosis of diabetes mellitus was supplemented by the use of any anti-diabetic medications, or, wherever available, any HbA1c level >6.5% (24); the diagnosis of hypertension was supplemented by the use of any anti-hypertensives; and the diagnosis of chronic kidney disease was supplemented, wherever available, by a baseline estimated glomerular filtration rate of <60 mL/min/1.73m^2^ as calculated from the 2021 Chronic Kidney Disease Epidemiology Collaboration (CKD-EPI) formula (25,26).

### Statistical analysis

VVLV was measured by the coefficient of variation (CV; defined by 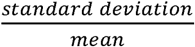). Continuous variables were expressed as mean ± standard deviation (SD). Due to the nature of the database and since patients with missing lipid levels were excluded, there were no missing values in this study. The association between the mean, SD, and CV of lipid measures and the risk of outcomes was tested using Cox regression, with hazard ratio (HR) and the corresponding 95% confidence intervals (CI) as summary statistics. Four separate Cox models were fitted hierarchically for the primary outcome. The first model was an unadjusted, univariable model including only the mean, SD, or CV of lipid measures. The second model was adjusted for age, sex, and the number of respective lipid tests. On top of the second model, the third model was further adjusted for comorbid conditions as listed above in *Data collection and definitions*. Finally, on top of the third model, the fourth model was further adjusted for medication prescriptions as listed above in *Data collection and definitions*. Variability measures were analysed as standardized continuous variables, such that the HR reflects increases in hazard per SD increase in the variability measure. To further illustrate the association of VVLV with the risk of the primary outcome, the CV of each lipid were also analysed in terciles using the fully adjusted Cox model (fourth model), for which the respective cumulative freedom from events was visualized using Kaplan-Meier curves.

Two *a priori* subgroup analyses were performed for the primary outcome using the fully adjusted multivariable Cox model with variability measures as continuous variables: the first by sex, and the second by any use of statins/fibrates at baseline. Two *a priori* sensitivity analyses were also performed for the primary outcome using the fully adjusted multivariable Cox model with variability measures as continuous variables. First, as mortality may constitute a competing event to the primary outcome and thus bias Cox regression findings, multivariable competing risk regression using the Fine and Gray sub-distribution model was performed, with death from any cause as the competing event and adjusting for the same variables as specified for the fourth Cox model above; sub-hazard ratio (SHR) and the corresponding 95% CI were used as summary statistics. Second, as cancer is a chronic disease that requires time to develop, inadequate follow-up duration may bias findings. The second sensitivity analysis thus consisted of fitting the fourth Cox model on only patients with at least two years of follow-up.

All p-values were two-sided, with p≤0.05 considered statistically significant. All analyses were performed on Stata v16.1 (StataCorp LLC, College Station, Texas, United States of America).

## Results

In total, 155,065 patients were identified by the inclusion criteria. After applying the exclusion criteria (**Figure 1**), 69,186 patients were included in the final analysis (26,679 male (38.6%); mean age 60.1±12.5 years old). The included patients had an average of 12.5±7.3 LDL-C tests, 12.6±7.3 HDL-C tests, 14.1±8.4 total cholesterol tests, and 14.3±8.5 triglycerides tests. The baseline characteristics of included patients are summarized in **Table 1**.

**Figure 1.**
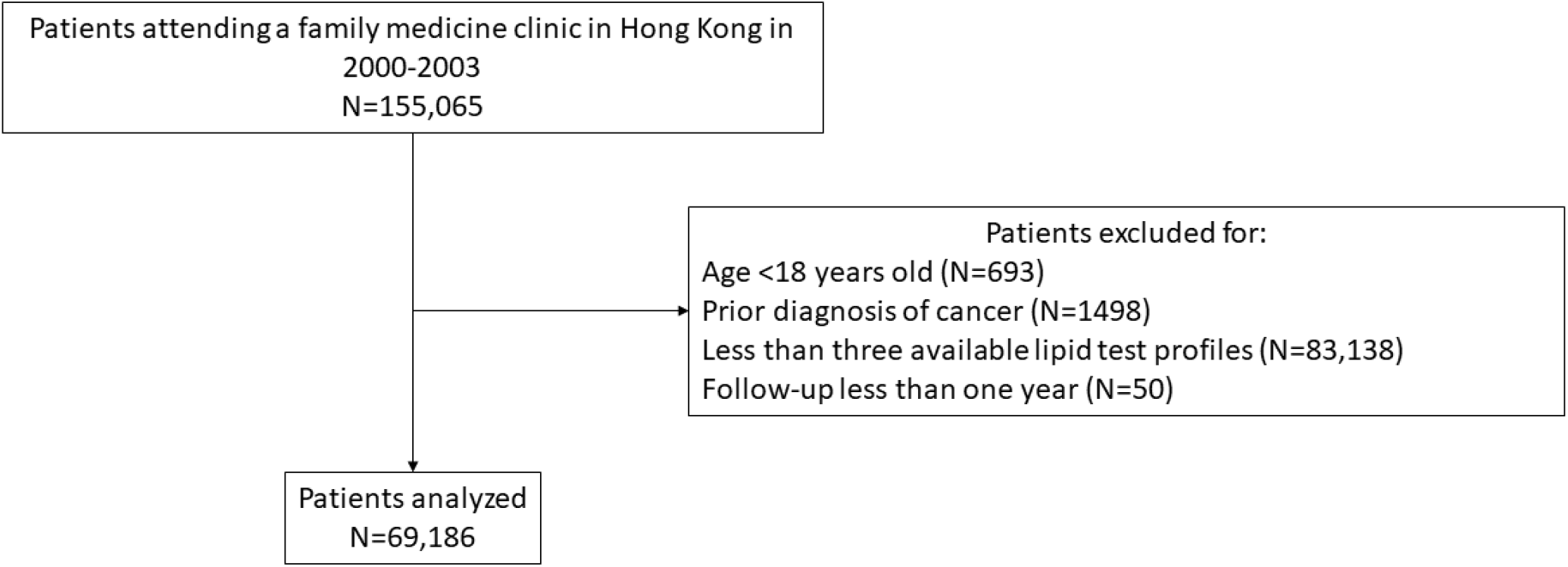
Study flowchart.

**Table 1.** Baseline characteristics of included patients.

| | Count (%) or mean ± standard deviation |
| --- | --- |
| Follow-up duration, years | 16.3±3.0 |
| Age, years old | 60.1±12.5 |
| Male, N (%) | 26,679 (38.6) |
| Hypertension, N (%) | 45,023 (65.1) |
| Heart failure, N (%) | 323 (0.5) |
| Atrial fibrillation, N (%) | 348 (0.5) |
| Myocardial infarction, N (%) | 152 (0.2) |
| Ischaemic heart disease, N (%) | 1360 (2.0) |
| Peripheral vascular disease, N (%) | 61 (0.1) |
| Stroke or transient ischaemic attack, N (%) | 448 (0.7) |
| Diabetes mellitus, N (%) | 20,263 (29.3) |
| Dyslipidaemia, N (%) | 16,206 (23.4) |
| Chronic kidney disease, N (%) | 6465 (9.3) |
| ACEI/ARB user, N (%) | 8395 (12.1) |
| Beta-blocker user, N (%) | 14,330 (20.7) |
| Calcium channel blocker user, N (%) | 14,134 (20.4) |
| Statin / fibrate user, N (%) | 8221 (11.9) |
| Antiplatelet user, N (%) | 6928 (10.0) |
| Metformin user, N (%) | 4989 (7.2) |
| Sulphonylurea user, N (%) | 5529 (8.0) |
| Insulin user, N (%) | 580 (0.8) |
| Mean LDL-C, mmol/L | 2.73±0.67 |
| CV of LDL-C | 0.22±0.11 |
| Number of LDL-C tests | 12.5±7.3 |
| Mean HDL-C, mmol/L | 1.36±0.35 |
| CV of HDL-C | 0.12±0.06 |
| Number of HDL-C tests | 12.6±7.3 |
| Mean total cholesterol, mmol/L | 4.78±0.78 |
| CV of total cholesterol | 0.14±0.07 |
| Number of total cholesterol tests | 14.1±8.4 |
| Mean triglycerides, mmol/L | 1.47±0.72 |
| CV of triglycerides | 27.4±13.6 |
| Number of triglycerides tests | 14.3±8.5 |

ACEI, angiotensin-converting enzyme inhibitor. ARB, angiotensin receptor blocker. CV, coefficient of variation. HDL-C, high-density lipoprotein cholesterol. LDL-C, low-density lipoprotein cholesterol.

Over a mean follow-up duration of 16.3±3.0 years, there were 7958 cases (11.5%) of incident cancer, with an incidence rate of 7.27 [95% confidence interval: 7.11, 7.43] cases per 1000 person-year. During the study duration, 14,091 patients (20.4%) died without developing any cancer.

### Association between lipid measures and the risk of incident cancer

Table 2 summarizes results of the Cox regression models evaluating the association between lipid measures and the risk of incident cancer. In the fully adjusted model, higher VVLV of all assessed types of lipids, as reflected by the CV, were associated with significantly higher risk of incident cancer (HR for LDL-C: 1.04 [1.02, 1.07], p<0.001; HR for HDL-C: 1.13 [1.11, 1.15], p<0.001; HR for total cholesterol: 1.05 [1.03, 1.08], p<0.001; HR for triglycerides: 1.04 [1.02, 1.07], p=0.001). Consistently, patients in the highest tercile of variability in LDL-C (HR 1.06 [1.00, 1.12], p=0.049; **Figure 2A**), HDL-C (HR 1.37 [1.29, 1.44], p<0.001; **Figure 2B**), total cholesterol (HR 1.10 [1.04, 1.17], p=0.001; **Figure 2C**) and triglycerides (HR 1.11 [1.06, 1.18], p<0.001; **Figure 2D**) had significantly elevated risk of incident cancer compared to those in the lowest tercile.

**Table 2.**
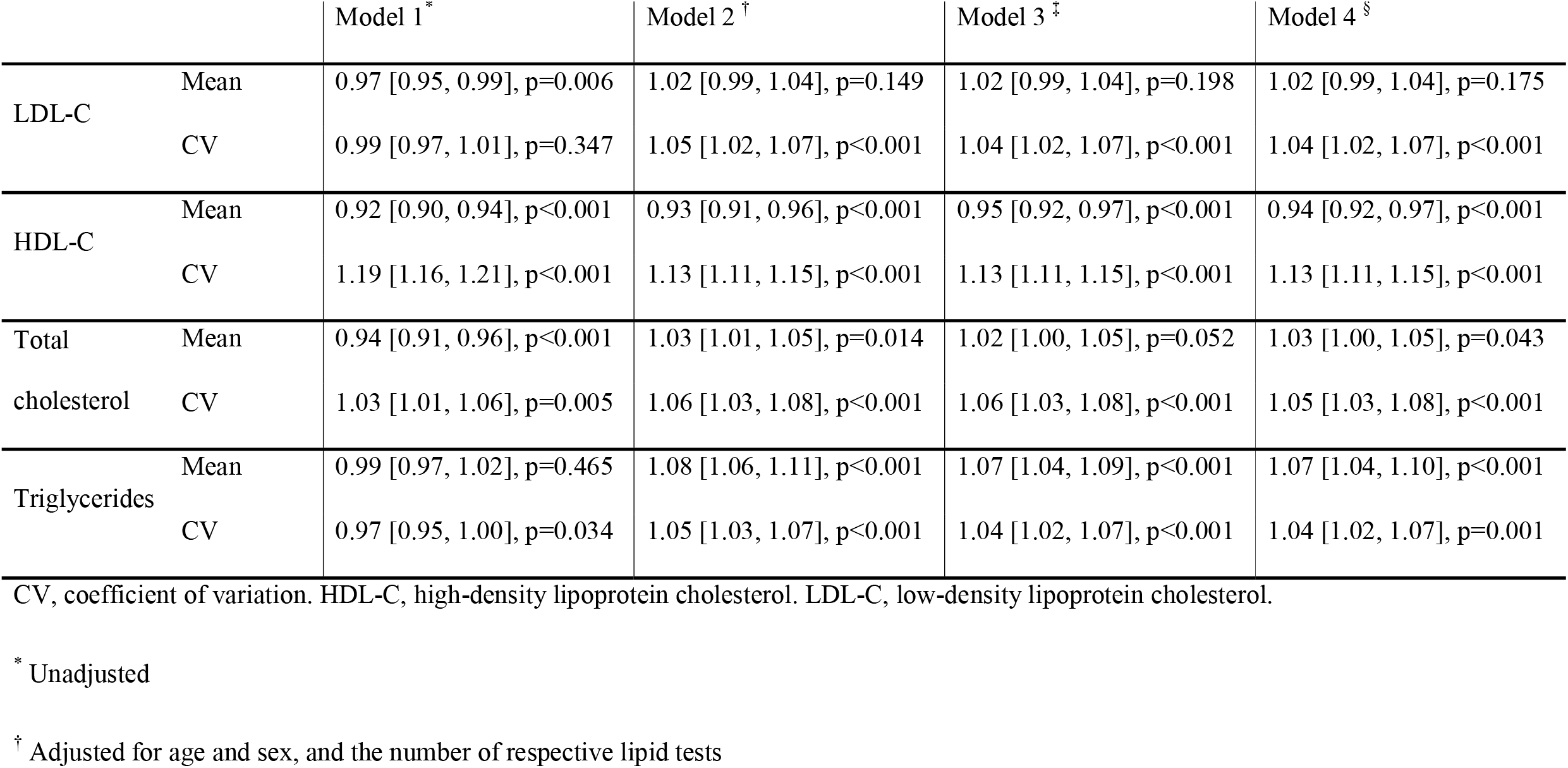
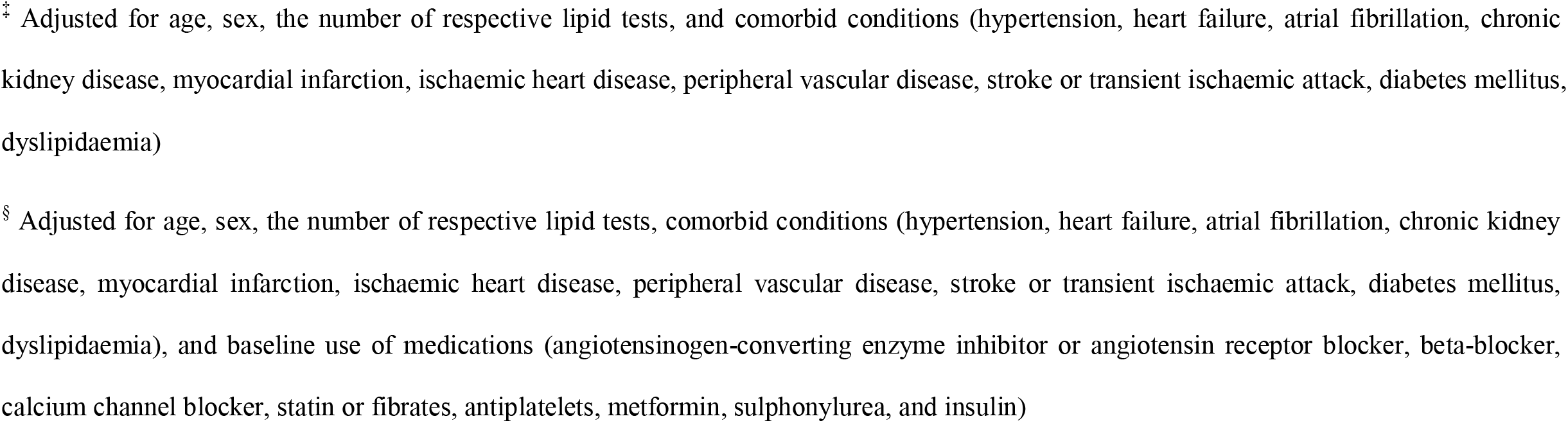
Association between lipid measures and the risk of incident cancer. Hazard ratios with the corresponding 95% confidence intervals are shown. All hazard ratios are per standard deviation increase in the measure.

|  |  | Model 1 * | Model 2 † | Model 3 ‡ | Model 4 § |
| --- | --- | --- | --- | --- | --- |
| LDL-C | Mean | 0.97 [0.95, 0.99], p=0.006 | 1.02 [0.99, 1.04], p=0.149 | 1.02 [0.99, 1.04], p=0.198 | 1.02 [0.99, 1.04], p=0.175 |
|  | CV | 0.99 [0.97, 1.01], p=0.347 | 1.05 [1.02, 1.07], p<0.001 | 1.04 [1.02, 1.07], p<0.001 | 1.04 [1.02, 1.07], p<0.001 |
| HDL-C | Mean | 0.92 [0.90, 0.94], p<0.001 | 0.93 [0.91, 0.96], p<0.001 | 0.95 [0.92, 0.97], p<0.001 | 0.94 [0.92, 0.97], p<0.001 |
|  | CV | 1.19 [1.16, 1.21], p<0.001 | 1.13 [1.11, 1.15], p<0.001 | 1.13 [1.11, 1.15], p<0.001 | 1.13 [1.11, 1.15], p<0.001 |
| Total cholesterol | Mean | 0.94 [0.91, 0.96], p<0.001 | 1.03 [1.01, 1.05], p=0.014 | 1.02 [1.00, 1.05], p=0.052 | 1.03 [1.00, 1.05], p=0.043 |
|  | CV | 1.03 [1.01, 1.06], p=0.005 | 1.06 [1.03, 1.08], p<0.001 | 1.06 [1.03, 1.08], p<0.001 | 1.05 [1.03, 1.08], p<0.001 |
| Triglycerides | Mean | 0.99 [0.97, 1.02], p=0.465 | 1.08 [1.06, 1.11], p<0.001 | 1.07 [1.04, 1.09], p<0.001 | 1.07 [1.04, 1.10], p<0.001 |
|  | CV | 0.97 [0.95, 1.00], p=0.034 | 1.05 [1.03, 1.07], p<0.001 | 1.04 [1.02, 1.07], p<0.001 | 1.04 [1.02, 1.07], p=0.001 |

CV, coefficient of variation. HDL-C, high-density lipoprotein cholesterol. LDL-C, low-density lipoprotein cholesterol.
\* Unadjusted
† Adjusted for age and sex, and the number of respective lipid tests
‡ Adjusted for age, sex, the number of respective lipid tests, and comorbid conditions (hypertension, heart failure, atrial fibrillation, chronic kidney disease, myocardial infarction, ischaemic heart disease, peripheral vascular disease, stroke or transient ischaemic attack, diabetes mellitus, dyslipidaemia)
§ Adjusted for age, sex, the number of respective lipid tests, comorbid conditions (hypertension, heart failure, atrial fibrillation, chronic kidney disease, myocardial infarction, ischaemic heart disease, peripheral vascular disease, stroke or transient ischaemic attack, diabetes mellitus, dyslipidaemia), and baseline use of medications (angiotensinogen-converting enzyme inhibitor or angiotensin receptor blocker, beta-blocker, calcium channel blocker, statin or fibrates, antiplatelets, metformin, sulphonylurea, and insulin)

**Figure 2.**
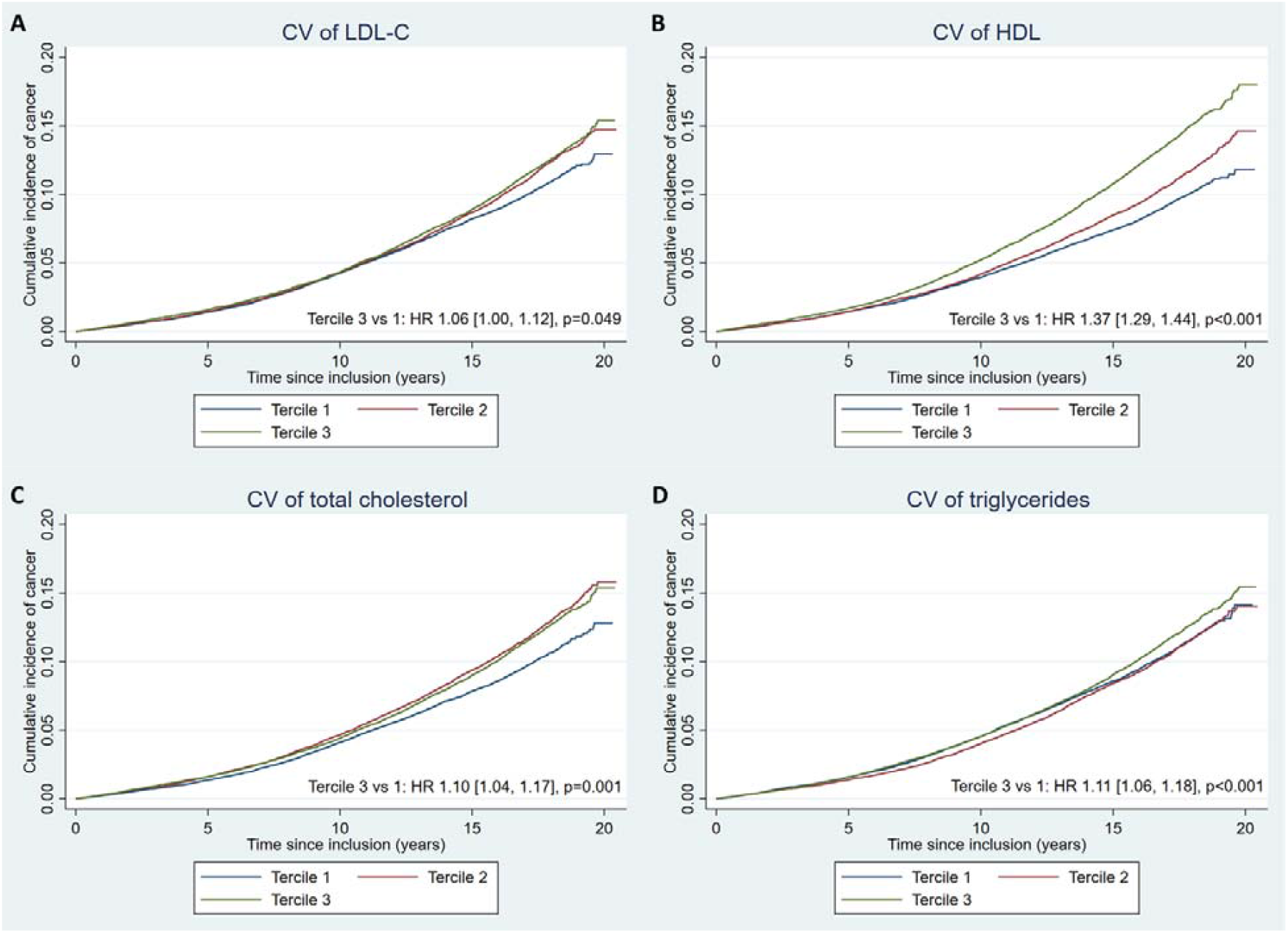
Kaplan-Meier incidence curves showing the cumulative incidence of cancer for patients in each tercile of the coefficient of variation (CV) for (**A**) low-density lipoprotein cholesterol (LDL-C), (**B**) high-density lipoprotein cholesterol (HDL-C), (**C**) total cholesterol, and (**D**) triglycerides. Hazard ratios (HR) shown are fully adjusted for covariates.

Meanwhile, lower mean HDL-C, and higher mean total cholesterol and mean triglycerides were associated with significantly higher risk of incident cancer; mean LDL-C, however, was not associated with the risk of incident cancer.

### Subgroup analyses

Results of the subgroup analysis by sex are summarized in **Supplementary Table 2**. The associations between lipid measures and the risk of incident cancer were qualitatively similar for both sexes, except for variability in triglycerides which was significantly associated with the risk of incident cancer in males (HR 1.07 [1.03, 1.10], p<0.001), but not females (HR 1.03 [1.00, 1.07], p=0.085). Additionally, the association between higher HDL-C variability and the risk of incident cancer was significantly stronger in males (HR for males: 1.17 [1.14, 1.20], p<0.001; HR for females: 1.10 [1.07, 1.13], p<0.001; p value for interaction <0.001). Significant interactions were also observed for mean HDL-C (p value for interaction 0.033) and mean triglycerides (p value for interaction 0.007), both of which showed a significantly stronger association with the risk of incident cancer in females.

Results of the subgroup analysis by use of statins/fibrates are summarized in **Supplementary Table 3**. Variability in triglycerides interacted significantly with the use of statins/fibrates (p value for interaction 0.024), with higher variability in triglycerides being associated with significantly higher risk of incident cancer in patients who did not use statins/fibrates (HR 1.05 [1.02, 1.08], p<0.001), but not in those who used these medications (HR 0.97 [0.91, 1.03], p=0.280). Similarly, the interaction between variability in LDL-C and the use of statins/fibrates approached but did not reach statistical significance (p value for interaction 0.084), with higher variability in LDL-C being associated with significantly higher risk of incident cancer in patients who did not use statins/fibrates (HR 1.05 [1.02, 1.08], p<0.001), but not in those who used these medications (HR 0.99 [0.93, 1.05], p=0.694). Additionally, lower mean HDL-C was associated with higher risk of incident cancer in patients who did not use statins/fibrates (HR 0.94 [0.92, 0.97], p<0.001), but not those who used these medications (HR 0.98 [0.92, 1.05], p=0.648); the interaction was statistically insignificant (p value for interaction 0.344) despite the qualitative difference. All other lipid measures’ associations with the risk of incident cancer were qualitatively consistent between subgroups without any significant interaction.

### Sensitivity analyses

Findings of the multivariable competing risk regression using the Fine and Gray subdistribution model (**Supplementary Table 4**) were largely similar to those of multivariable Cox regression, except for mean total cholesterol which, in this sensitivity analysis, was not significantly associated with the incidence of incident cancer (SHR 1.01 [0.98, 1.03], p=0.537). Findings of the sensitivity analysis including only patients with at least two years of follow-up (N=69,095; **Supplementary Table 5**) were also similar to those of the main analysis, with the association for mean total cholesterol marginally remaining statistically significant (HR 1.02 [1.00, 1.05], p=0.050).

## Discussion

In this population-based cohort study of 69,186 Chinese adults with a mean follow up of more than 15 years, we demonstrated that higher visit-to-visit variability of LDL-C, HDL-C, TC, and TG levels was associated with a significantly increased risk of incident cancer. Additionally, we found that lower mean HDL-C, and higher mean TC and mean TG were associated with significantly higher risk of incident cancer. The statistical significance of these associations was maintained under sensitivity analyses using the Fine and Gray sub-distribution model and various stratified analyses. To the best of our knowledge, the present study is one of the first to report on the association between VVLV and the risk of incident cancer.

### Potential Underlying Mechanisms

While the exact mechanisms underlying the observed associations between VVLV and incident cancer are unclear, several potential explanations exist. A potential mechanism that is immediately apparent would be a dysfunctional lipid metabolism. Lipid metabolism is known to be a critical component of carcinogenesis, regulating the synthesis and thus activity of multiple signalling pathways that are directly carcinogenic, or those that lead to pro-tumorigenic environments (27). An important example of the latter is a pro-inflammatory state, which, in turn, has been observed in patients with elevated VVLV. Such association has been shown by several prior studies (15,28), and was indirectly supported by possible associations between VVLV and endothelial dysfunction (29,30), a marker of chronic inflammation (31,32). As inflammation plays a critical role in the cancer development and progression (33–35), higher VVLV may reflect an underlying proinflammatory state, which in turn mediate the observed association between VVLV and elevated risk of incident cancer.

The known links between VVLV and incident cardiovascular diseases may also shed light on other possible mechanisms underlying the observed associations (11–14). Cancer and many cardiovascular diseases, such as heart failure, share additional pathophysiological mechanisms including neuro-hormonal activation, oxidative stress, and a dysfunctional immune system (36). It is thus possible that these overlapping mechanisms may have mediated the associations observed in this study. Nonetheless, it is important to note that whilst our findings demonstrated association between VVLV and the risk of incident cancer, causality was not investigated. Specifically, reverse causation remains possible, where developing cancer may increase VVLV before cancer diagnosis, resulting in a spurious association between VVLV and the risk of incident cancer.

### Comparison with previous literature

Few studies had explored the relationship between VVLV and the risk of incident cancer. One such study was performed by Choi and colleagues, who studied a cohort of Korean adults over a median follow-up duration of 5.1 years and showed that higher HDL-C variability were associated with an increased risk of multiple myeloma (37). In contrast to the limited scope of the study by Choi and colleagues, who focused solely on multiple myeloma, we showed that higher VVLV was broadly associated with the risk of incident cancer. Results of the current study further support the clinical relevance of VVLV by showing that all lipid variability parameters are important predictors for incident cancer. In addition, our study had a substantially longer follow-up duration that exceeded 15 years on average. Altogether, findings of this study and that by Choi and colleagues strongly suggested VVLV as a clinically relevant marker for incident cancer, and may signal new opportunities to further our understanding of the roles played by lipid metabolism in cancer development.

### Clinical Implications and the future

Our findings suggest that VVLV may be considered as a potential tool for cancer risk assessment. We derived these findings from a cohort of family medicine patients, who essentially represent the least unwell patients that one would encounter in daily clinical practice. Given the ubiquity and low cost of lipid testing, VVLV has the potential to achieve widespread clinical use with minimal interference of general medical practices. This potentially kills two birds with one stone, as including VVLV in general medical assessments may allow not only stratification of cancer risk, but also cardiovascular risk (11–14). Higher VVLV may alert clinicians for closer and more intensive monitoring, follow-up, and workup in these regards. At the minimum, we hope that our findings raise clinicians’ awareness of the importance of VVLV and reduce the likelihood of clinically important variability being dismissed as mere measurement errors.

Nevertheless, much remains to be done before widespread clinical utilization of VVLV can become a reality, including determination of clinical thresholds for decision-making, and further investigation of effect modifiers that may influence the prognostic power of VVLV. A case in point for the latter would be the use of lipid-lowering medications such as statins and fibrates, which we observed to have negated some of the associations between VVLV and the risk of incident cancer. This was consistent with a recent study, in which the use of lipid-lowering medications was observed to partially negate the association between VVLV and the risk of incident heart failure (11).

Furthermore, VVLV may also be a novel modifiable risk factor for cancer. Our findings should therefore encourage further research into interventions, pharmacological or behavioural alike, that optimize lipid variability. Recently, there has been increasingly robust evidence supporting the use of statins in cancer prevention (38–40). Given the known effects of statins on lipid levels, as well as the aforementioned interactions between statin use and the prognostic power of VVLV, VVLV may be further explored as both a mediator and a treatment target for patients using statins. Our findings may also prompt further investigations in the prognostic value of VVLV for different, specific types of cancers, for which there is no existing evidence to the best of our knowledge, with the single exception of the aforementioned study on multiple myeloma by Choi and colleagues (37).

### Strengths and Limitations

The present study included a large number of patients from a population-based using data from routine clinical practice. As the database covers an estimated 90% of Hong Kong’s population (17), the patient data analysed in this study may be considered as representative of real-world practice in Hong Kong, and is likely generalizable to other Asian regions. Moreover, the robustness of the analyses were reinforced by the use of multiple sensitivity analyses, in which similar observations were made consistently.

Nonetheless, our study is not free of limitations. First, given the observational nature of the study, there is inherent information bias due to under-coding, coding errors, and missing data. Nevertheless, previous studies have demonstrated good coding accuracy and data completeness in CDARS (41), and CDARS has been used extensively in research (18–22). Although individual adjudication of patient data was not possible due to the deidentified nature of the database, all the data was entered by treating clinicians without any involvement from any of the authors, and none of the authors had the right or authority to alter the data. Second, there might be unmeasured and residual confounders that we have not accounted for, which may include other comorbidities, smoking status, body composition, or intercurrent illnesses. Nonetheless, we have adjusted for a range of well-established risk factors in the multivariable Cox regression analyses, which should account for most potential confounding factors pertinent to our outcomes. Third, CDARS only recorded prescription data, but not medication adherence. Thus, the possible effects of medication non-adherence on lipid variability and the prognostic values of these measures of variability could not be estimated. Nonetheless, our subgroup analyses showed substantial differences between subgroups, suggesting that usage of lipid-lowering medication(s) indeed had significant effects on the variability measures’ prognostic values.

In conclusion, higher VVLV was associated with an increased risk of incident cancer in the general population. Some of these associations might be negated by the use of lipid-lowering medications such as statins and fibrates. However, further research is warranted to evaluate the role of VVLV in relationship with specific cancer subtypes, as well as the underlying pathophysiological mechanisms driving these associations.

## Data Availability

All data were available on reasonable request to the corresponding author.

## Institutional review board approval

This study has been approved by the Joint Chinese University of Hong Kong– New Territories East Cluster Clinical Research Ethics Committee

