## Supplementary Table for "Association between visit-to-visit lipid variability and incident cancer: a population-based cohort study"

**Supplementary Material**

**Supplementary Table 1.** ICD-9 codes used to identify outcomes and co-morbidities.

| Heart failure | 428 428.1 428.2 428.2 428.21 428.22 428.23 428.3 428.3 428.31 428.32 428.33 428.4 428.4 428.41 428.42 428.43 428.9 398.91 402.01 402.11 402.91 404.01 404.03 404.11 404.13 404.91 404.93 |
| --- | --- |
| Myocardial infarction | 410.01 410.02 410.1 410.11 410.12 410.2 410.21 410.22 410.3 410.31 410.32 410.4 410.41 410.42 410.5 410.51 410.52 410.6 410.61 410.62 410.7 410.71 410.72 410.8 410.81 410.82 410.9 410.91 410.92 |
| Diabetes mellitus | 250 250.01 250.02 250.03 250.1 250.11 250.12 250.13 250.2 250.21 250.22 250.23 250.3 250.31 250.32 250.33 250.4 250.41 250.42 250.43 250.5 250.51 250.52 250.53 250.6 250.61 250.62 250.63 250.7 250.71 250.72 250.73 250.8 250.81 250.82 250.83 250.9 250.91 250.92 250.93 |
| Hypertension | 401 401.1 401.9 402 402.01 402.1 402.11 402.9 402.91 403 403.01 403.1 403.11 403.9 403.91 404 404.01 404.02 404.03 404.1 404.11 404.12 404.13 404.9 404.91 404.92 404.93 405 405.01 405.09 405.1 405.11 405.19 405.9 405.91 405.99 437.2 |
| Atrial fibrillation | 427.31 429.4 |
| Stroke / transient ischaemic attack | 435 435.1 435.2 435.3 435.8 435.9 433.81 433.91 434 436 437 437.1 433.31 433.01 434.01 434.1 434.11 434.9 434.91 437.2 437.3 437.4 437.5 437.6 437.7 437.8 437.9 430 431 432 432.1 432.9 |
| Peripheral vascular disease | 250.7 443.9 443 443.1 443.2 443.21 443.22 443.23 443.24 443.29 443.8 443.81 443.82 443.89 441 443.9 785.4 V43.4 |
| Dyslipidaemia | 242.0 242.1 242.2 242.3 242.4 |
| Ischaemic heart disease | 410.01 410.02 410.1 410.11 410.12 410.2 410.21 410.22 410.3 410.31 410.32 410.4 410.41 410.42 410.5 410.51 410.52 410.6 410.61 410.62 410.7 410.71 410.72 410.8 410.81 410.82 410.9 410.91 410.92 411 411.1 411.8 411.81 411.89 413 413.1 413.9 414 414.01 414.02 414.03 414.04 414.05 414.06 414.07 414.1 414.11 414.12 414.19 414.2 414.3 414.4 414.8 414.9 410 412 |
| Cancer | 140 140.1 140.3 140.4 140.5 140.6 140.8 140.9 141 141.1 141.2 141.3 141.4 141.5 141.6 141.8 141.9 142 142.1 142.2 142.8 142.9 143 143.1 143.8 143.9 144 144.1 144.8 144.9 145 145.1 145.2 145.3 145.4 145.5 145.6 145.8 145.9 146 146.1 146.2 146.3 146.4 146.5 146.6 146.7 146.8 146.9 147 147.1 147.2 147.3 147.8 147.9 148 148.1 148.2 148.3 148.8 148.9 149 149.1 149.8 149.9 150 150.1 150.2 150.3 150.4 150.5 150.8 150.9 151 151.1 151.2 151.3 151.4 151.5 151.6 151.8 151.9 152 152.1 152.2 152.3 152.8 152.9 153 153.1 153.2 153.3 153.4 153.5 153.6 153.7 153.8 153.9 154 154.1 154.2 154.3 154.8 155 155.1 155.2 156 156.1 156.2 156.8 156.9 157 157.1 157.2 157.3 157.4 157.8 157.9 158 158.8 158.9 159 159.1 159.8 159.9 160 160.1 160.2 160.3 160.4 160.5 160.8 160.9 161 161.1 161.2 161.3 161.8 161.9 162 162.2 162.3 162.4 162.5 162.8 162.9 163 163.1 163.8 163.9 164 164.1 164.2 164.3 164.8 164.9 165 165.8 165.9 170 170.1 170.2 170.3 170.4 170.5 170.6 170.7 170.8 170.9 171 171.2 171.3 171.4 171.5 171.6 171.7 171.8 171.9 172 172.1 172.2 172.3 172.4 172.5 172.6 172.7 172.8 172.9 173 173.01 173.02 173.09 173.1 173.11 173.12 173.19 173.2 173.21 173.22 173.29 173.3 173.31 173.32 173.39 173.4 173.41 173.42 173.49 173.5 173.51 173.52 173.59 173.6 173.61 173.62 173.69 173.7 173.71 173.72 173.79 173.8 173.81 173.82 173.89 173.9 173.91 173.92 173.99 174 174.1 174.2 174.3 174.4 174.5 174.6 174.8 174.9 175 175.9 176 176.1 176.2 176.3 176.4 176.5 176.8 176.9 179 180 180.1 180.8 180.9 181 182 182.1 182.8 183 183.2 183.3 183.4 183.5 183.8 183.9 184 184.1 184.2 184.3 184.4 184.8 184.9 185 186 186.9 187 187.1 187.2 187.3 187.4 187.5 187.6 187.7 187.8 187.9 188 188.1 188.2 188.3 188.4 188.5 188.6 188.7 188.8 188.9 189 189.1 189.2 189.3 189.4 189.8 189.9 190 190.1 190.2 190.3 190.4 190.5 190.6 190.7 190.8 190.9 191 191.1 191.2 191.3 191.4 191.5 191.6 191.7 191.8 191.9 192 192.1 192.2 192.3 192.8 192.9 193 194 194.1 194.3 194.4 194.5 194.6 194.8 194.9 195 195.1 195.2 195.3 195.4 195.5 195.8 200 200.01 200.02 200.03 200.04 200.05 200.06 200.07 200.08 200.1 200.11 200.12 200.13 200.14 200.15 200.16 200.17 200.18 200.2 200.21 200.22 200.23 200.24 200.25 200.26 200.27 200.28 200.3 200.31 200.32 200.33 200.34 200.35 200.36 200.37 200.38 200.4 200.41 200.42 200.43 200.44 200.45 200.46 200.47 200.48 200.5 200.51 200.52 200.53 200.54 200.55 200.56 200.57 200.58 200.6 200.61 200.62 200.63 200.64 200.65 200.66 200.67 200.68 200.7 200.71 200.72 200.73 200.74 200.75 200.76 200.77 200.78 200.8 200.81 200.82 200.83 200.84 200.85 200.86 200.87 200.88 201 201.01 201.02 201.03 201.04 201.05 201.06 201.07 201.08 201.1 201.11 201.12 201.13 201.14 201.15 201.16 201.17 201.18 201.2 201.21 201.22 201.23 201.24 201.25 201.26 201.27 201.28 201.4 201.41 201.42 201.43 201.44 201.45 201.46 201.47 201.48 201.5 201.51 201.52 201.53 201.54 201.55 201.56 201.57 201.58 201.6 201.61 201.62 201.63 201.64 201.65 201.66 201.67 201.68 201.7 201.71 201.72 201.73 201.74 201.75 201.76 201.77 201.78 201.9 201.91 201.92 201.93 201.94 201.95 201.96 201.97 201.98 202 202.01 202.02 202.03 202.04 202.05 202.06 202.07 202.08 202.1 202.11 202.12 202.13 202.14 202.15 202.16 202.17 202.18 202.2 202.21 202.22 202.23 202.24 202.25 202.26 202.27 202.28 202.3 202.31 202.32 202.33 202.34 202.35 202.36 202.37 202.38 202.4 202.41 202.42 202.43 202.44 202.45 202.46 202.47 202.48 202.5 202.51 202.52 202.53 202.54 202.55 202.56 202.57 202.58 202.6 202.61 202.62 202.63 202.64 202.65 202.66 202.67 202.68 202.7 202.71 202.72 202.73 202.74 202.75 202.76 202.77 202.78 202.8 202.81 202.82 202.83 202.84 202.85 202.86 202.87 202.88 202.9 202.91 202.92 202.93 202.94 202.95 202.96 202.97 202.98 203 203.01 203.02 203.1 203.11 203.12 203.8 203.81 203.82 204 204.01 204.02 204.1 204.11 204.12 204.2 204.21 204.22 204.8 204.81 204.82 204.9 204.91 204.92 205 205.01 205.02 205.1 205.11 205.12 205.2 205.21 205.22 205.3 205.31 205.32 205.8 205.81 205.82 205.9 205.91 205.92 206 206.01 206.02 206.1 206.11 206.12 206.2 206.21 206.22 206.8 206.81 206.82 206.9 206.91 206.92 207 207.01 207.02 207.1 207.11 207.12 207.2 207.21 207.22 207.8 207.81 207.82 208 208.01 208.02 208.1 208.11 208.12 208.2 208.21 208.22 208.8 208.81 208.82 208.9 208.91 208.92 196 196.1 196.2 196.3 196.5 196.6 196.8 196.9 197 197.1 197.2 197.3 197.4 197.5 197.6 197.7 197.8 198 198.1 198.2 198.3 198.4 198.5 198.6 198.7 198.8 198.81 198.82 198.89 199 199.1 |

**Supplementary Table 2.** Subgroup analyses by sex with the fully adjusted multivariable Cox regression model.

|  | |  | Hazard ratio [95% confidence interval] | p value for interaction |
| --- | --- | --- | --- | --- |
| LDL-C | Mean | Male  Female | 1.03 [0.99, 1.07], p=0.118  1.00 [0.97, 1.04], p=0.847 | 0.728 |
|  | CV | Male  Female | 1.04 [1.00, 1.07], p=0.045  1.05 [1.02, 1.09], p=0.001 | 0.232 |
| HDL-C | Mean | Male  Female | 0.96 [0.92, 1.00], p=0.031  0.92 [0.89, 0.95], p<0.001 | 0.033 |
|  | CV | Male  Female | 1.17 [1.14, 1.20], p<0.001  1.10 [1.07, 1.13], p<0.001 | <0.001 |
| Total cholesterol | Mean | Male  Female | 1.02 [0.98, 1.06], p=0.277  0.99 [0.96, 1.02], p=0.600 | 0.800 |
|  | CV | Male  Female | 1.08 [1.05, 1.12], p<0.001  1.05 [1.02, 1.09], p=0.001 | 0.373 |
| Triglycerides | Mean | Male  Female | 1.06 [1.02, 1.11], p=0.002  1.10 [1.06, 1.14], p<0.001 | 0.007 |
|  | CV | Male  Female | 1.07 [1.03, 1.10], p<0.001  1.03 [1.00, 1.07], p=0.085 | 0.905 |

CV, coefficient of variation. HDL-C, high-density lipoprotein cholesterol. LDL-C, low-density lipoprotein cholesterol.

**Supplementary Table 3.** Subgroup analyses by use of statins/fibrates with the fully adjusted multivariable Cox regression model.

|  | |  | Hazard ratio [95% confidence interval] | p value for interaction |
| --- | --- | --- | --- | --- |
| LDL-C | Mean | Users  Non-users | 1.04 [0.98, 1.11], p=0.198  1.01 [0.99, 1.04], p=0.317 | 0.367 |
|  | CV | Users  Non-users | 0.99 [0.93, 1.05], p=0.694  1.05 [1.02, 1.08], p<0.001 | 0.084 |
| HDL-C | Mean | Users  Non-users | 0.98 [0.92, 1.05], p=0.648  0.94 [0.92, 0.97], p<0.001 | 0.344 |
|  | CV | Users  Non-users | 1.12 [1.06, 1.19], p<0.001  1.13 [1.11, 1.15], p<0.001 | 0.574 |
| Total cholesterol | Mean | Users  Non-users | 1.05 [0.98, 1.12], p=0.165  1.00 [0.97, 1.02], p=0.884 | 0.146 |
|  | CV | Users  Non-users | 1.02 [0.95, 1.09], p=0.604  1.07 [1.04, 1.10], p<0.001 | 0.156 |
| Triglycerides | Mean | Users  Non-users | 1.06 [1.00, 1.11], p=0.046  1.06 [1.03, 1.09], p<0.001 | 0.815 |
|  | CV | Users  Non-users | 0.97 [0.91, 1.03], p=0.280  1.05 [1.02, 1.08], p<0.001 | 0.024 |

CV, coefficient of variation. HDL-C, high-density lipoprotein cholesterol. LDL-C, low-density lipoprotein cholesterol.

**Supplementary Table 4.** Competing risk analysis with full multivariable adjustments using the Fine and Gray sub-distribution model.

|  | | Sub-hazard ratio [95% confidence interval] |
| --- | --- | --- |
| LDL-C | Mean | 1.00 [0.98, 1.02], p=0.992 |
|  | CV | 1.04 [1.02, 1.06], p=0.001 |
| HDL-C | Mean | 0.97 [0.94, 0.99], p=0.005 |
|  | CV | 1.10 [1.07, 1.12], p<0.001 |
| Total cholesterol | Mean | 1.01 [0.98, 1.03], p=0.537 |
|  | CV | 1.04 [1.01, 1.06], p=0.003 |
| Triglycerides | Mean | 1.04 [1.01, 1.07], p=0.002 |
|  | CV | 1.03 [1.00, 1.05], p=0.027 |

CV, coefficient of variation. HDL-C, high-density lipoprotein cholesterol. LDL-C, low-density lipoprotein cholesterol.

**Supplementary Table 5.** Sensitivity analysis of patients with at least two years of follow-up (N=69,095).

|  | | Hazard ratio [95% confidence interval] |
| --- | --- | --- |
| LDL-C | Mean | 1.02 [0.99, 1.04], p=0.200 |
|  | CV | 1.04 [1.02, 1.07], p<0.001 |
| HDL-C | Mean | 0.94 [0.92, 0.97], p<0.001 |
|  | CV | 1.13 [1.11, 1.15], p<0.001 |
| Total cholesterol | Mean | 1.02 [1.00, 1.05], p=0.050 |
|  | CV | 1.05 [1.03, 1.08], p<0.001 |
| Triglycerides | Mean | 1.07 [1.04, 1.10], p<0.001 |
|  | CV | 1.04 [1.02, 1.07], p=0.001 |

CV, coefficient of variation. HDL-C, high-density lipoprotein cholesterol. LDL-C, low-density lipoprotein cholesterol.
